# Exploration of Short-range Neonatal Seizure Forecasting with Quantitative EEG Based Deep Learning

**DOI:** 10.1101/2023.08.01.23293285

**Authors:** Jonathan Kim, Hannah C Glass, Edilberto Amorim, Vikram R Rao, Danilo Bernardo

## Abstract

**Background:** In this study, we utilize robust feature selection of quantitative encephalography (QEEG) features for inclusion into a deep learning (DL) model for short-range forecasting of neonatal seizure risk.

**Methods:** We used publicly available EEG seizure datasets with a total of 132 neonates. The Boruta algorithm with Shapley values was used for QEEG feature selection into a convolutional long short-term memory (ConvLSTM) DL model to classify preictal versus interictal states. ConvLSTM was trained and evaluated with 10-fold cross-validation. Performance was evaluated with varying seizure prediction horizons (SPH) and seizure occurrence periods (SOP).

**Results:** Boruta with Shapley values identified statistical moments, spectral power distributions, and RQA features as robust predictors of preictal states. ConvLSTM performed best with SPH 3 min and SOP 7 min, demonstrating 80% sensitivity with 36% of time spent in false alarm, AUROC of 0.80, and AUPRC of 0.23. The model demonstrated ECE of 0.106, consistent with moderate calibration. Evaluation of forecasting skill with BSS under varying SPH demonstrated a peak BSS of 0.056 and a trend for decreasing BSS with increasing SPH.

**Conclusion:** Statistical moments, spectral power, and recurrence quantitative analysis are predictive of the preictal state. Short-range neonatal seizure forecasting is feasible with DL models utilizing these features.

## Introduction

Neonatal seizures, with an incidence rate of one to three per 1000 life births, are associated with substantial long-term morbidity and mortality (1, 2). Prompt seizure treatment is critical for neonates, as a higher seizure burden is associated with increased treatment resistance and mortality (3, 4, 5, 6). A promising strategy to improve neonatal clinical outcomes has focused on identifying seizure-prone neonates using clinical and EEG features to reduce the time to seizure diagnosis and treatment (7, 8, 9, 10, 11).

Recent studies have leveraged machine learning (ML) to predict seizures in neonatal encephalopathy (NE) with high accuracy utilizing long forecast horizons during the acute postnatal period (10, 11). Pavel et al. introduced a neonatal ML model utilizing clinical variables and quantitative EEG (QEEG) features shortly after birth to predict neonates with NE who later developed seizures, forecasting individual seizure risk over several days (10). Recently, McKee et al. developed a ML model on qualitative EEG and clinical features from the first day of life that could predict subsequent seizures during the acute monitoring period spanning days (11). While these and other prior studies have predicted seizure risk over an observation period spanning several days (7, 8, 9, 10, 11), short-range seizure risk forecasting, or forecasts with higher temporal resolution, remains unexplored in the neonatal population.

In contrast to long-range forecasts, short-range forecasting provides more precise and timely information regarding the imminence of seizure onset (Figure 1a). The provision of short-range forecasts may facilitate the investigation of prophylactic interventions in higher seizure risk populations such as NE, and help optimize the allocation of monitoring resources, which in many environments is limited in accessibility to continuous EEG (12, 13, 14). Thus, our objective was to extend upon prior work in neonatal seizure forecasting, by investigating high-temporal-resolution forecasting spanning minutes.

**Figure 1.**
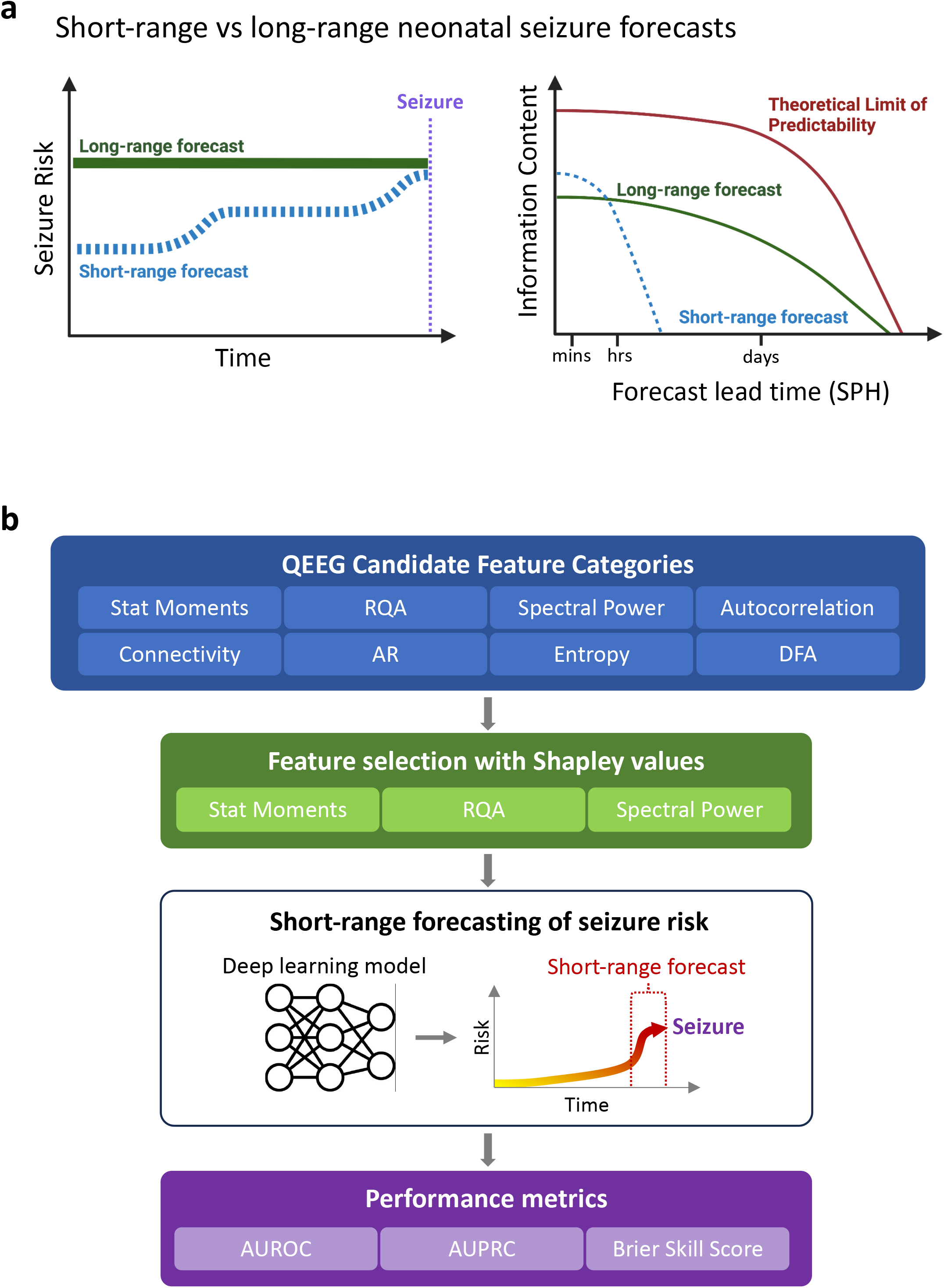
a. The left panel illustrates differences between short-and long-range forecasting. Short-range forecasts may be updated frequently (indicated by dynamics in the dashed line), which enables these forecasts to be revised regularly as new data becomes available. Current neonatal long-range seizure forecasting typically provide time-invariant, or static forecasts. The right panel illustrates the relation between forecast information content and forecasting lead time, following analogous forecasting definitions previously developed in meteorology (41). Here, *information content* may pertain to the degree of accuracy, certainty, and overall applicability of the forecasts. There is a downward trend in information content in the theoretical limit of predictability, short-and long-range forecast curves reflecting that in a complex and chaotic systems such as the brain, there is an inevitable increase in uncertainty as one attempts to forecasts further into the future with increasing lead time, or SPH. The short-range forecast may potentially contain higher information content than long-range forecasts during short lead times because it is based on more up to date observations. However, as the SPH becomes longer, for both short-and long-range forecasts, the information content decreases due to increased uncertainty and a reduction in the accuracy of forecasts. The long-range forecast curve starts after one hour after birth reflecting current long-range forecasting models that are based on information obtained from at least the first hour of EEG information. b. Starting from a broad selection of features across different categories, feature selection utilizing the Boruta algorithm utilizing Shapley value feature importances was performed to identify features most predictive of interictal and preictal states. Feature categories in which had significant importance were incorporated into ConvLSTM to estimate time-varying, short-range seizure risk. Feature Abbreviations: Recurrence quantification analysis (RQA), Detrended Fluctuation Analysis (DFA), Autoregressive Modeling (AR), Statistical Moments (Stat Moments) Metric Abbreviations: Area Under Receiver Operator Characteristic Curve (AUROC), Area Under Precision Recall Curve (AUPRC)

Here, we develop a deep learning (DL) approach for QEEG-based neonatal seizure forecasting and utilize publicly available neonatal EEG datasets to evaluate model performance on short-range forecasting of neonatal seizures. Considering the uncertainty into which specific QEEG features are most predictive of the preictal state in neonates, we integrated robust QEEG feature selection methods into our approach.

## Methods

### Subject Data

We utilized two publicly available EEG datasets from Helsinki University Hospital (HUH) and Cork University Maternity Hospital (Cork). The HUH dataset consists of multi-channel 256 Hz EEG recorded from 79 term neonates at the NICU in HUH, Helsinki, Finland, with total 60 hours of recording (15). In the HUH dataset, the presence of seizures in the EEGs was annotated independently by three experts. The most common diagnosis in this dataset was birth asphyxia (35 patients). The Cork dataset from the INFANT Research Center, Cork University Maternity Hospital, contains EEG records from 53 neonates affected by HIE with total 169 hours total recording (16). While the majority of HUH subjects contained ictal samples, only two Cork subjects have EEG records containing seizures in this dataset, thus providing relatively more balance between seizure-containing and non-seizure containing subjects.

### Preprocessing

We segmented EEG data into 20-second non-overlapping epochs with class labels of preictal, interictal, and ictal states. *Ictal periods* were defined as time periods in which at least 2 experts annotated a seizure. We defined *preictal periods* as between 6 min to 1 min prior to seizure onset and *interictal periods* as between 1 minute after end of seizure to 5 min prior to the next seizure. Right-censored periods in which it is unknown whether a seizure occurred within the prediction window at the end of data recordings were excluded. Prior to windowing and feature calculation, the raw EEG signal was band-pass filtered between 0.1 Hz and 20 Hz, and then resampled from the original 256 Hz to two times the Nyquist frequency, 40 Hz. For seizures that are less than the seizure prediction horizon (SPH) from the previous seizure, we consider them as a single seizure event. To increase model robustness and generalizability, we performed augmentation of the training dataset by translation invariant transforms; specifically, we transposed the raw EEG channels across the frontal-occipital and left-right axes. Candidate QEEG features from feature categories were calculated on non-overlapping 20-second EEG epochs.

### Feature Selection

Feature selection refers to the process of identifying the subset of features most pertinent to a prediction model’s performance. Unlike automatic or emergent feature extraction methods, such as those used by convolutional neural networks, QEEG features may vary significantly in relevance and predictive power, necessitating a robust feature selection process. Increasing the number of features without bounds, or excessively high-dimensional QEEG data, may impair model performance due to increased data complexity, a concept broadly known in neuroscience and other domains as the ’curse of dimensionality’ (17). Overfitting also becomes a concern, as models trained on a multitude of features may undermine generalizability to novel data. Feature selection, distilling the feature set down to those of true relevance, enhances model generalization, streamlines model training, and improves model interpretation (18).

For feature selection, we utilized BorutaSHAP(19), which integrates the robustness of the Boruta algorithm feature selection strategy with the Shapley value feature importances derived from SHapley Additive exPlanations (20). The Boruta algorithm is a feature selection method used in ML, which is based on the random forest classification algorithm(21). It utilizes feature importances, such as SHAP or Gini, to measure to identify significantly predictive features in a dataset. These feature importances are iteratively compared with those of shadow features, which are generated from random shuffling of the real features to provide a reference. A threshold for feature selection is defined by the maximum importance score derived from the shadow features. Using this threshold, two-sided T-test is used to ascertain the relative significance of each feature—features significantly below the threshold are considered ’unimportant’, while those significantly above the threshold are deemed ’important’. This feature importance ranking utilizes Shapley values, a game-theoretic method that determines individual feature contributions to model predictions, which provides consistent, accurate feature importance scores. Shapley values represent each feature’s average marginal contribution to model prediction, across all possible combinations(20). Through this integrated BorutaSHAP procedure, we identified the top feature categories consistently surpassing the threshold, thereby indicating significant correlation with the preictal or interictal state.

Selected QEEG features from the top three feature categories are demonstrated in Supplementary Table 1 in more detail, which included standard summary statistics (mean, standard deviation, kurtosis, skew, the 10th percentile, and the 90th percentile), calculated across each montaged channel left-right pair, and power spectral features calculated at each montage channel (22). Other feature categories previously utilized for seizure prediction in prior studies that were evaluated included autocorrelation, entropy, detrended fluctuation analysis, and coherence (23).

### Model Design

We developed a custom convolutional long short-term memory neural network (ConvLSTM), an architecture that has previously been utilized for seizure prediction (24). An advantage of utilizing an LSTM-based architecture relative to conventional ML methods is their capability to learn underlying temporal dependencies from sequential data. The incorporation of the convolutional layer allows for local temporal feature extraction. Further details regarding ConvLSTM architecture are shown in Supplementary Figure 1.

### Model Training

As described above, the data used to train the model consisted of three classes, preictal, inter-ictal, and ictal. The loss function used to train the model was formulated as a multi-class cross-entropy function that disregarded performance on the ictal class while penalizing mislabeling those members of preictal and interictal classes. The pytorch and pytorch-lightning libraries were used to evaluate models(25). Grid-search optimization was used to tune ConvLSTM and demonstrated best-performance with a learning rate of 5e-5, a maximum of 100 training epochs, and early stopping of training conditioned on validation loss delta of 1e-6. For each fold of the 10-fold cross-validation, the entire dataset was split into one train/validation set consisting of 90% of the data and one test set consisting of 10% of the data. The train/validation set was then further split into a training set and a validation set, with the training set taking 75% of the train/validation set and the validation set taking the remaining 25%. All splits were created with inter-subject stratification, ensuring that data from a given subject was either solely in the train set or solely in the test set.

### Model Performance Evaluation

We employed the seizure alarm framework, which utilizes the seizure prediction horizon (SPH) and seizure occurrence period (SOP) (26). In this framework, a minimum SPH is designated to ensure sufficient lead time preceding a seizure to allow for timely intervention strategies to be employed. Concurrently, a maximum SOP is set to the desired temporal resolution suited to the duration of applicability of a given forecast. If the predicted risk is above the designated threshold, an alarm is triggered, lasting the combined duration of SPH and SOP. Details regarding the definitions of SPH and SOP are shown in Supplementary Figure 2. We evaluated area under the ROC curve (AUROC), area under the precision-recall curve (AUPRC), Expected Calibration Error (ECE), and Brier-Skill Score (BSS). PR-AUC illustrates the trade-off between precision (the proportion of true positives among all predicted positives) and recall (the proportion of true positives among all actual positives). It is advantageous in imbalanced datasets such as in this case, where the number of interictal periods (negative class) significantly exceeds preictal periods (positive class), as it focuses on the model’s performance concerning the positive class. ECE is evaluated to measure the reliability of the model’s predicted probabilities and is commonly used to evaluate neural network performance (27). It assesses the discrepancy between the predicted and true probabilities of the outcomes, which is important in determining whether the model is well-calibrated. The BSS assesses the model’s forecasting skill relative to a reference random classifier model. The BSS considers both calibration and discrimination of the model. A positive BSS indicates that the model performs better than the reference model, while a negative BSS signifies the opposite. For the BSS calculation, we utilized the standard climatology reference, which accounts for the prevalence of the positive class (preictal states). For comparison of ConvLSTM to conventional ML methods, including Support Vector Machine (SVM), K-Nearest Neighbors (KNN), Logistic Regression, and random forest (RF) classifiers, we additionally evaluated F1 score and Matthew Correlation Coefficient (MCC).

## RESULTS

### Study Subjects

The HUH EEG dataset consists of 79 subjects with median age 40 weeks (interquartile range: 39.4 - 40.7) with 39 (49%) subjects with total of 516 across all subjects (28). The Cork EEG dataset consists of 53 subjects with median age 39.5 (37.8-40.5) weeks with two subjects (4%) who had seizures (16). The average duration per subjects of the HUH and Cork datasets were 1.2 (1.1-1.6) hrs and 3 (2–4) hours.

The study overview is demonstrated in Figure 1b, including all feature categories considered for feature selection. Feature selection with Boruta algorithm utilizing Shapley value feature importances demonstrated that the top three feature categories included statistical moments, spectral power, and RQA features (Figure 2). These features were identified as consistently demonstrating importance scores exceeding the threshold (Supplementary Figure 3), indicating their predictive relevance. QEEG features from the top three feature categories (Supplementary Table 1) were incorporated into ConvLSTM. Examples of the resulting time-varying seizure risk forecasts for individual neonates with seizures are shown in Figures 3. In these examples, periods containing heightened seizure risk occur with varying lead times prior to the seizure occurrences (red lines). In comparison, the selected examples from neonates with no seizures show no peaks indicative of preictal state (Figure 4).

**Figure 2.**
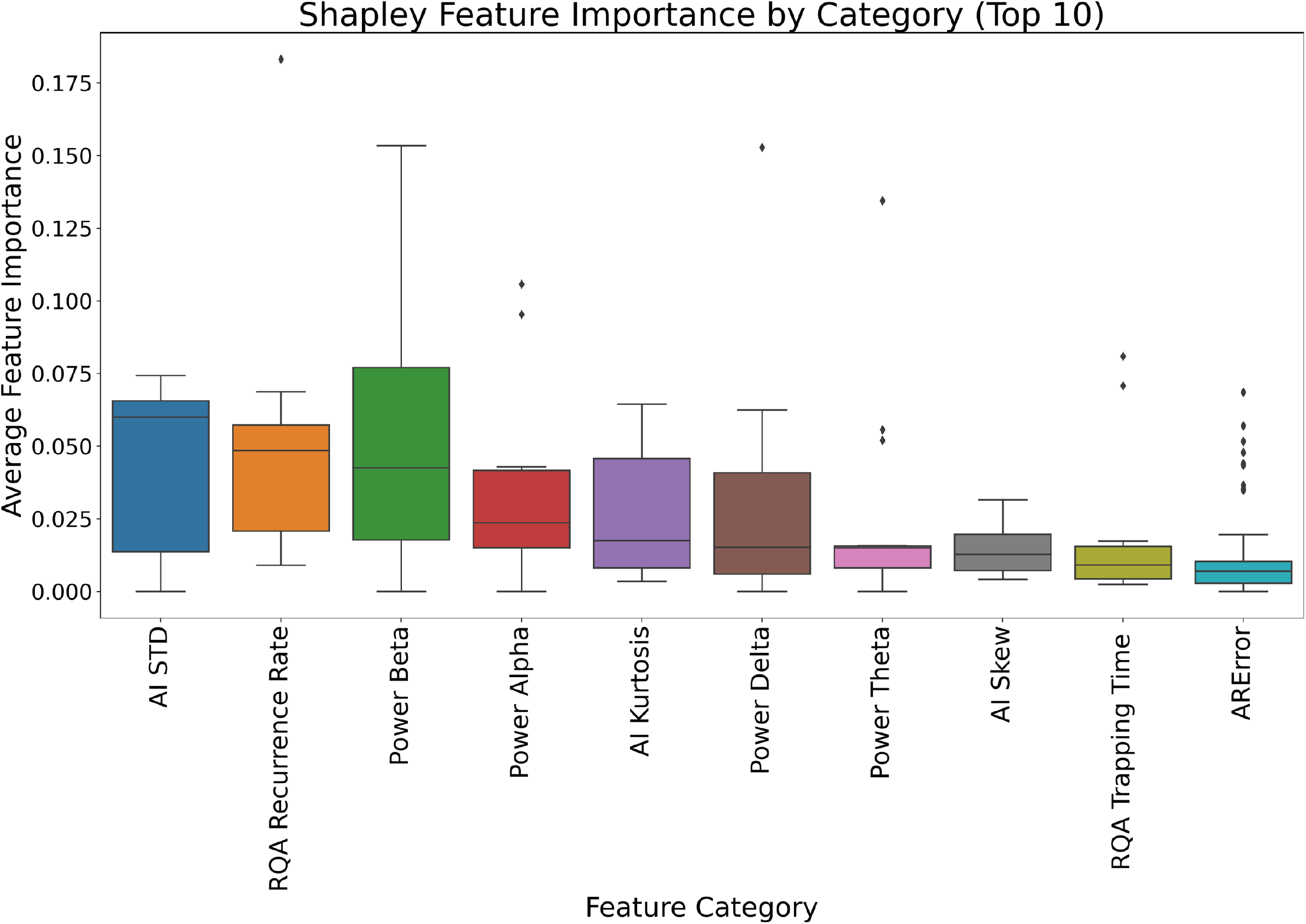
Boruta analysis was performed to identify the QEEG feature categories most predictive at classifying interictal and preictal states. The top feature categories included features from statistical moments (e.g. Standard deviation (STD) asymmetric index (AI)), spectral power distributions in different frequency bands, and RQA feature categories such as RQA recurrence rate. Features are ranked in accordance by their Shapley feature importance scores, which were utilized in the Boruta analysis. Abbreviations and formulations for features from the top 3 feature categories are defined in Supplementary Table 1.

**Figure 3.**
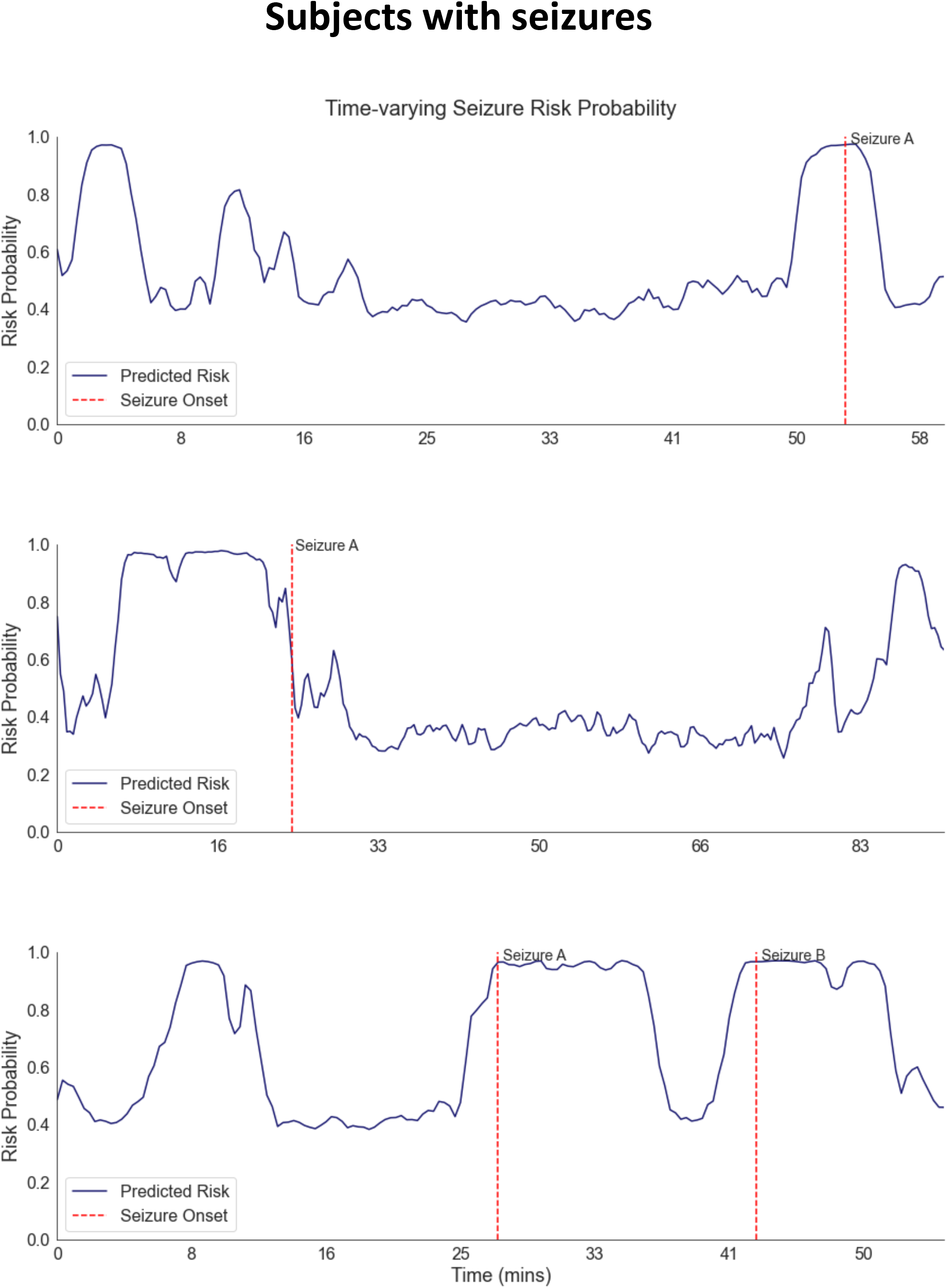
Examples from 3 subjects with seizures are shown. Prior to seizure occurrences, there are increases in estimated preictal state probability. There are also elevations in preictal state probability not associated with immediate seizure.

**Figure 4.**
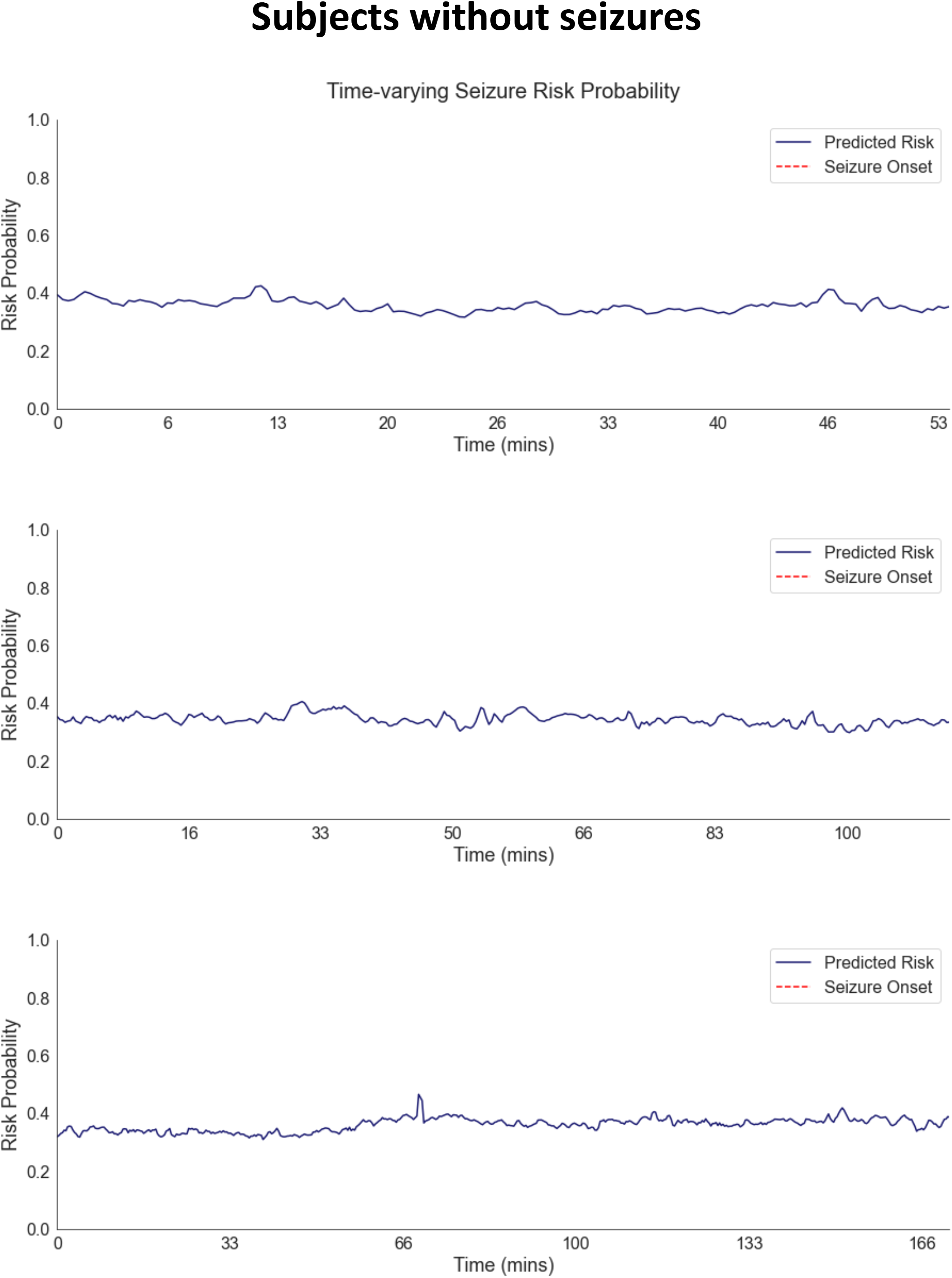
Examples from 3 subjects without seizures are shown. There are no significant elevations in preictal state probability seen.

To assess the performance of ConvLSTM over varying short-range forecast horizons, we evaluated area under the ROC curve and area under the AUPRC varying SOP and SPH between 1 to 7 minutes (Figure 5). Peak AUROC was 0.80 with SPH of 3 minutes and SOP of 7 minutes, and at these SPH and SOP settings, with ROC threshold adjusted to correspond with 80% sensitivity, there was a corresponding 36% time spent during false alarm. The peak AUPRC was 0.23 with SPH of 1 minute and SOP of 7 minutes. For any given SPH, increasing the SOP led to improved AUROC and AUPRC, reflective of increased ease of forecasting at lower temporal resolutions.

**Figure 5.**
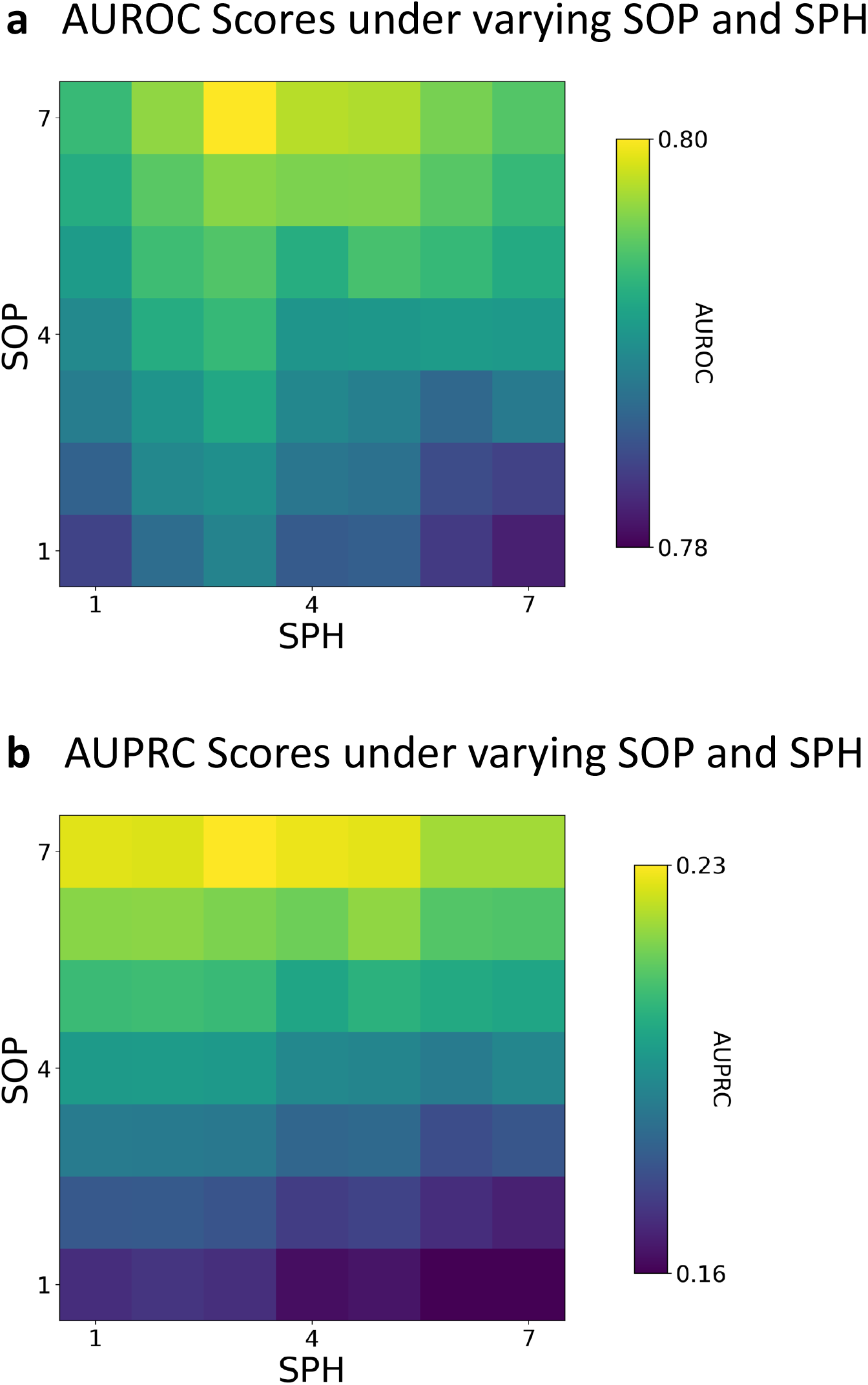
Area under the ROC and PR curve analyses are demonstrated for ConvLSTM model. A) shows respective AUROC’s for SOP and SPH varied between 1 and 7 minutes. The best AUROC was 0.80 at SOP of 7 minutes and SPH of 3 minutes. B) shows respective AUPRC’s for SOP and SPH varied between 1 and 7 minutes. The best AUPRC was 0.23 at SOP of 7 minutes and SPH of 3 minutes.

The calibration of the ConvLSTM model was evaluated using the reliability plot and Expected Calibration Error (ECE) metrics. The reliability plot, (Figure 6a), demonstrates the relationship between the predicted probabilities and the observed frequency of the preictal class. There is a clustering of points below the diagonal, which signifies that ConvLSTM was occasionally overconfident in its predictions for the preictal class at these probabilities. Additionally, isolated points distant from the diagonal occurred at the lowest and highest probability bins, consistent with respective isolated underconfident and overconfident forecasts at these bins. Concordant to the reliability plot findings, the ECE value of 0.106 is consistent with a moderately well-calibrated model. The value of 0.106 is slightly above the general desirable range for a strongly calibrated model, considered less than 0.1 (zero indicates perfect calibration, and one is the maximum value indicative of weak calibration).

**Figure 6.**
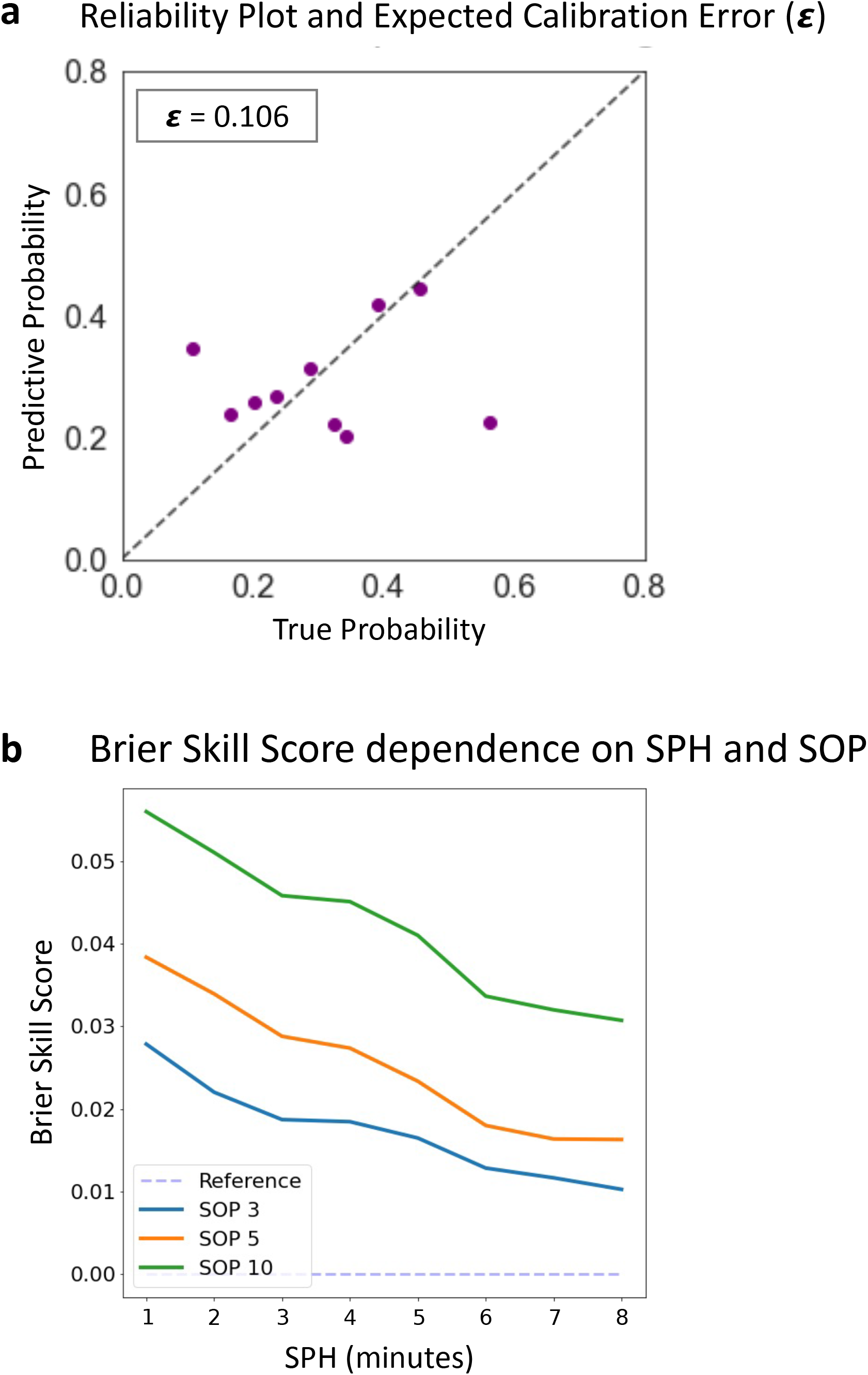
6a demonstrates ConvLSTM calibration evaluation using a reliability plot and Expected Calibration Error (ECE) metric. The ECE plot An ECE (χ) value of 0.106 indicates a moderately-well calibrated model. 6b demonstrates the influence of varying SPH and SOP on forecasting skill as evaluated by the Brier Skill Score (BSS). The best BSS occurred with SPH of 1 minute and SOP of 10 minutes. An increase in SPH correlates with a decline in BSS, and analogously, as the SOP decreases, model performance also decreases, indicating that forecasting with increasing lead-times and increasing temporal resolution becomes more challenging.

To evaluate the forecasting skill of ConvLSTM, we examined the impact of forecasting horizon and resolution utilizing the Brier Skill Score (BSS), which measures the difference between the accuracy of the model’s predictions and the accuracy of a reference forecast. The effect of varying SPH and SOP on BSS is shown in Figure 6b. The highest BSS was 0.056 obtained with a SPH of 1 minute and SOP of 10 minutes. We find that for all SOP, as the SPH increases, then model performance concomitantly decreases. This follows the intuition that forecasting farther into the future is inherently more difficult. Similarly, as the SOP temporal resolution becomes finer, model performance also decreases, suggesting that forecasting with increased temporal resolution is also more difficult.

Lastly, we compared ConvLSTM performance to conventional ML methods. ConvLSTM model achieved higher AUROC, AUPRC, and F1 scores than SVM, KNN, logistic regression, and random forest classifiers at the classification of preictal versus interictal states (Supplementary Table 2).

## DISCUSSION

In this study, we utilized the Boruta algorithm and Shapley values for robust selection of QEEG features, identifying statistical moments, spectral power, and RQA features as most predictive of interictal and preictal states. We incorporated these features into ConvLSTM and demonstrated the accuracy of providing short-term forecasts of neonatal seizures. Forecasting horizons as brief as 5 minutes has been reported in adult patients with long-term chronic intracranial EEG recordings (23, 29). However, to our knowledge, this is the first study to demonstrate short-range forecasting in the neonatal population. Short-range forecasting in the neonates at high risk of impending seizure may enable time-sensitive interventions in higher seizure risk populations, such as in neonatal encephalopathy, and help optimize the allocation of monitoring resources.

Notably, recent ML approaches at predicting neonatal seizure risk have utilized a combination of QEEG features with clinical features with ensemble ML methods such as gradient boosted decision trees and random forests to estimate seizure risk accurately (10, 11). In contrast to these prior works, which estimate neonatal seizure risk over several days, our approach focuses on short-term forecasting with a time resolution of minutes.

Recent seizure forecasting studies in the pediatric population have utilized subject-specific feature engineering and model training for their analyses (30, 31, 32, 33). Notably, Tsiouras et al. achieved 100% sensitivity with an FPR of 0.06/hr on the full CHB-MIT set by using a subject-specific neural network with a long short-term memory (LSTM) architecture (34). Our approach is distinct from these prior works in that we developed a subject-independent model instead of a subject-specific model. Although our performance metrics are moderately lower than those attained in prior patient-specific models, the subject-independent approach presented obviates the need for subject-specific training. This is particularly clinically advantageous for neonates who are often at immediate elevated risk of seizure after birth, such as in cases of neonatal encephalopathy, where subject-specific training data may not be immediately available before the first seizure. Furthermore, subject-independent models facilitate efficient resource allocation, as it can be readily implemented at different sites without fine-tuning, reducing the need for specialized expertise and computational resources.

Concerning our modeling approach, we utilized a convolutional LSTM that demonstrated overall improved performance compared to conventional ML methods. ConvLSTM incorporates strengths of both convolutional neural network (CNN) and LSTM architectures: the convolutional module can efficiently downsample the input signal while extracting local temporal features predictive of seizure risk, whereas the LSTM module effectively learns long-range temporal dependencies. Prior studies in seizure prediction have previously utilized CNN (33, 35, 36), LSTM (34, 37), and CNN-LSTM(24). In contrast to the previously published CNN-LSTM method, which utilized CNN-LSTM on short-time Fourier-transformed (STFT) EEG signal, we incorporated other QEEG features predictive of preictal states in addition to spectral power changes, including statistical moments and RQA features.

Regarding the limitations of our study, we recognize that we utilized a relatively low number of subjects and did not utilize a held-out or independent evaluation dataset. Thus, validation on larger and independent datasets are necessary to confirm our findings. Additionally, our model demonstrated a relatively higher false alarm rate and lower sensitivity than prior studies, which have predominantly utilized the CHB-MIT dataset (Supplementary Table 3), which may be attributed to our subject-independent approach. In addition, the relatively lower performance seen in this study may reflect the usage of shorter-range SOP and SPH, as forecasting with increasingly higher temporal resolution is considered more difficult in complex systems, such as in seismology or meteorology, and this has also been suggested for seizure forecasting (38, 39, 40).

In conclusion, we demonstrate the potential of applying ML approaches to enable time-dependent neonatal seizure forecasting, facilitating more precise timing and temporal understanding of neonatal seizure susceptibility.

## Data Availability

All data produced in the present study are available upon reasonable request to the authors.

---

**Supplementary Figure 1.**
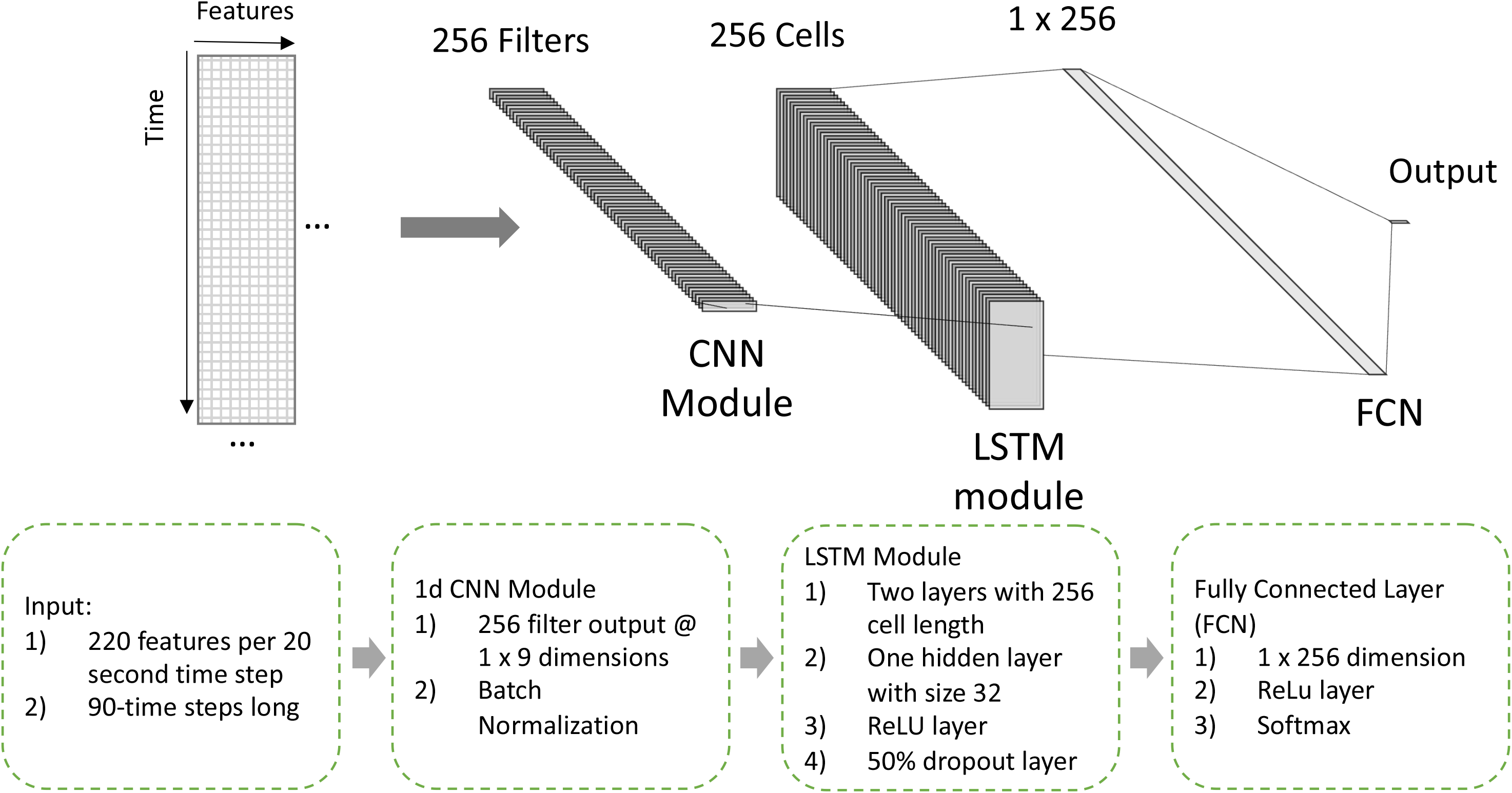
The ConvLSTM architecture consisted of a convolutional layer with 256 output filters, followed by a batch normalization layer, then followed by an LSTM module with 256 cells and one hidden layer with size of 32, followed by a rectified linear unit (ReLU), followed by a 50% dropout layer, a fully connected linear layer (FCN), and then a final ReLU layer as final output. A softmax function was applied to the final ReLU layer to yield probabilistic predictions for each output class. For visualization purposes only 64 filters/cells are shown as opposed to 256.

**Supplementary Figure 2.**
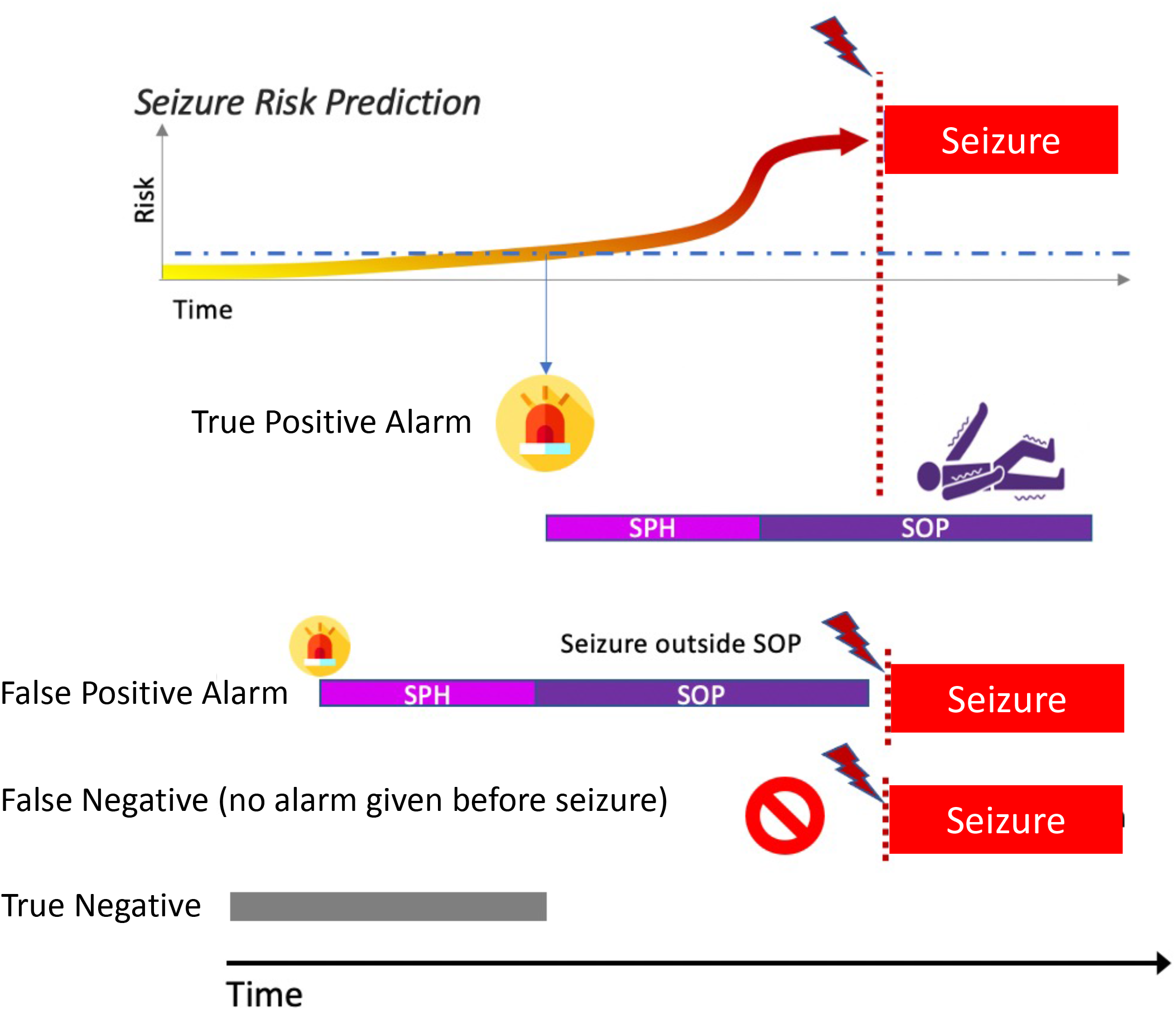
The seizure prediction horizon (SPH) and seizure occurrence period (SOP) evaluation framework considers that there should be a minimum SPH to provide ample lead time before a seizure to allow for intervention and that the alarm should have SOP selected to align forecast duration with the specified clinical observation period. The system triggers an alarm, lasting the combined duration of SPH and SOP, if the designated seizure threshold is met. At time *t*, a true positive alarm occurs if a seizure initiates between *t*+SPH and *t*+SPH+SOP; otherwise, a false positive is marked. A false negative occurs if a seizure occurs at time *t_s_* and no alarm is activated. A true negative occurs when no alarm is triggered and no seizure occurs.

**Supplementary Figure 3.**
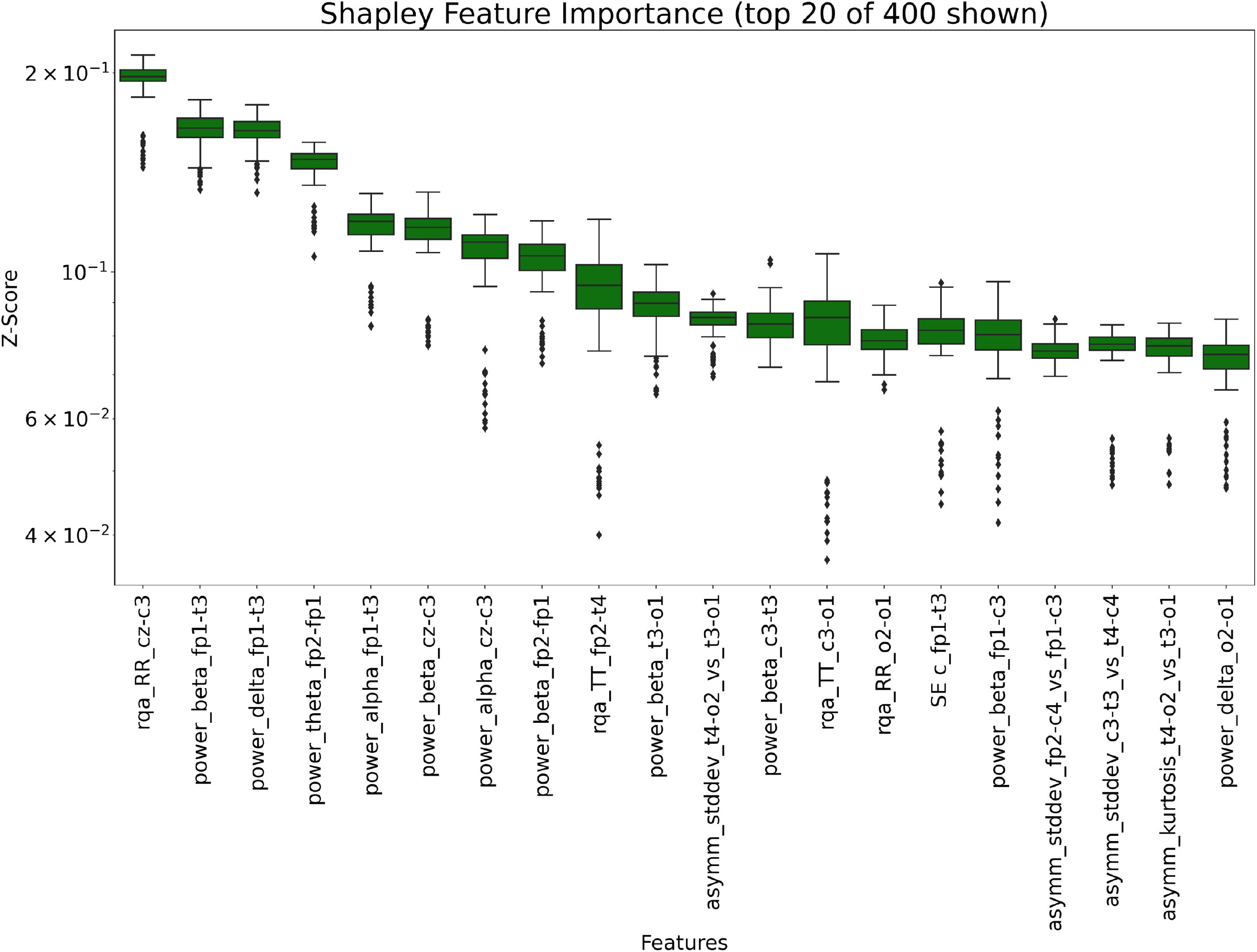
Boruta analysis was performed to identify the most predictive QEEG features at each channel. In the top 20 features the mean Shapley values were predominantly from statistical moments, spectral power distribution, and RQA feature categories. Abbreviations for these top 3 feature categories are defined in Supplementary Table 1.

## SUPPLEMENTARY TABLES

**Supplementary Table 1.**
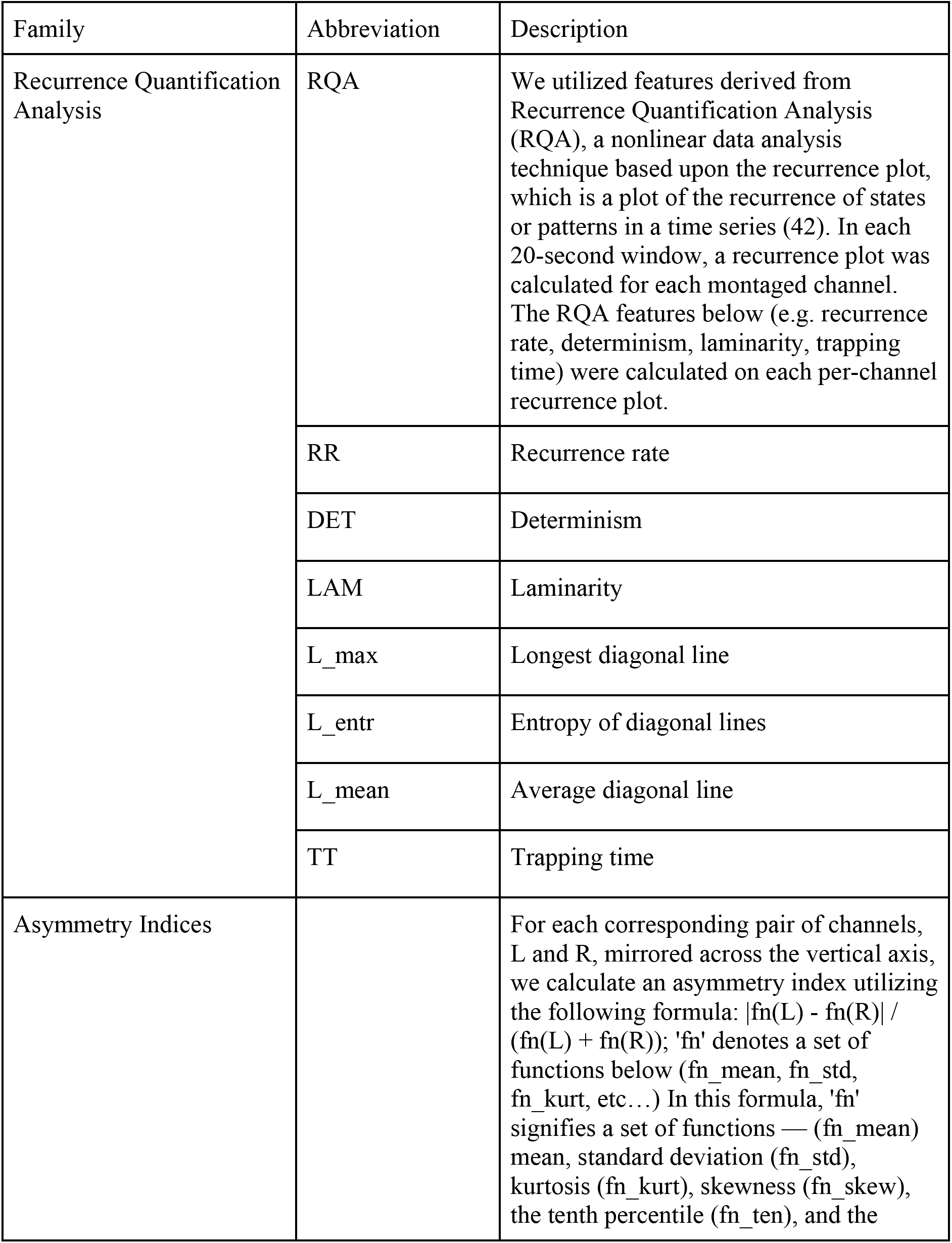
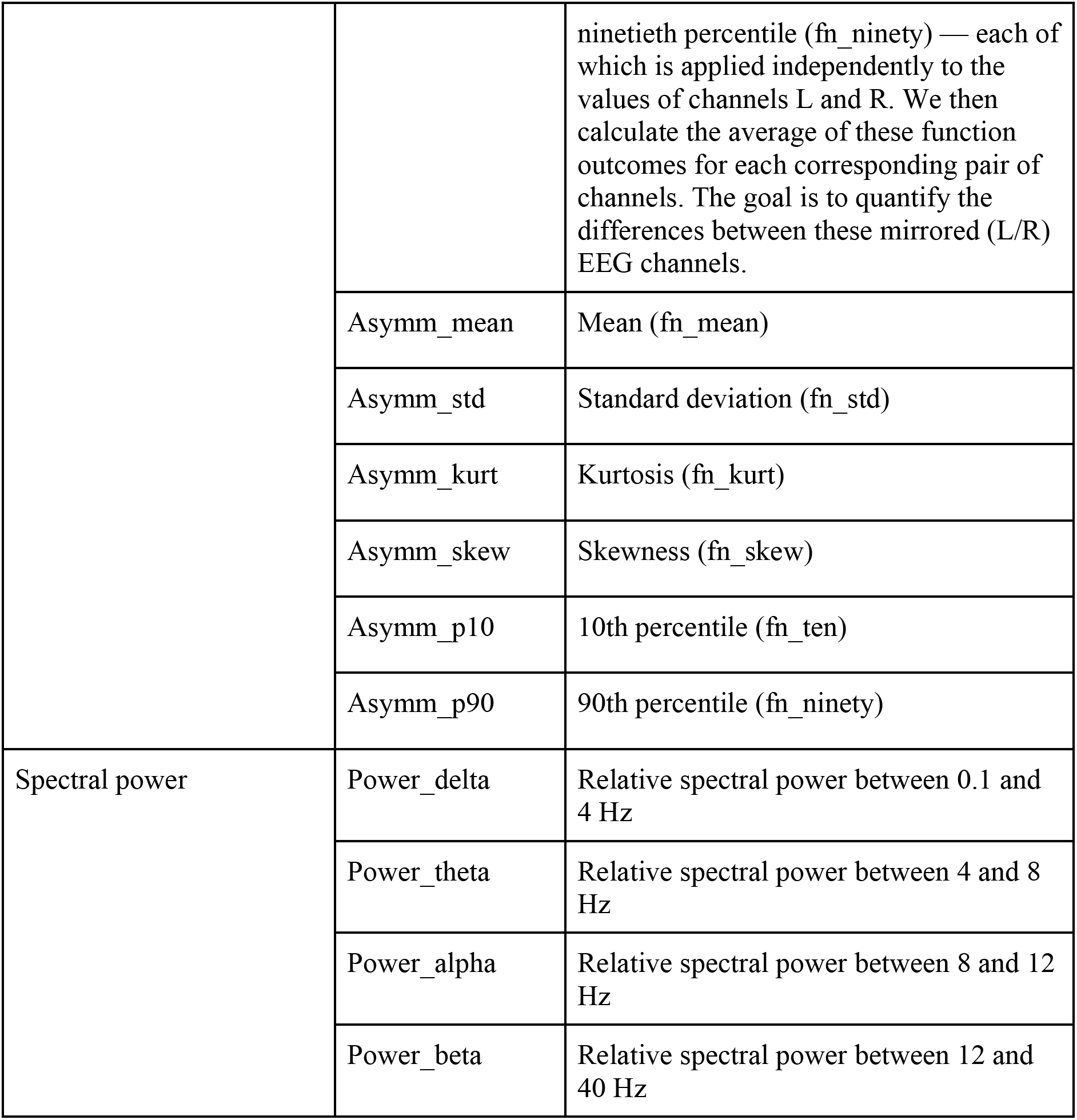

**Supplementary Table 2.**

|  | AUROC | AUPRC | MCC | F1 |
| --- | --- | --- | --- | --- |
| **ConvLSTM** | **0.678 ±0.041** | **0.218 ±0.059** | **0.255 ±0.054** | **0.334 ±0.050** |
| Random Forest | 0.612 ±0.021 | 0.147 ±0.023 | 0.141 ±0.024 | 0.252 ±0.030 |
| Support Vector Machine | 0.613 ±0.037 | 0.163 ±0.030 | 0.144 ±0.030 | 0.262 ±0.031 |
| Logistic Regression | 0.598 ±0.025 | 0.163 ±0.027 | 0.112 ±0.023 | 0.243 ±0.027 |
| K-Nearest Neighbors | 0.633 ±0.022 | 0.112 ±0.013 | 0.151 ±0.023 | 0.255 ±0.029 |

Data reported as average performance across all cross-validation folds (10) with ± standard error mean. Abbreviations: Matthew Correlation Coefficient (MCC). F1 score (F1).

**Supplementary Table 3.**

| Year | Authors | Dataset | Feats | CLF | Same Cal | # Sz | Sens (%) | FPR (/h) | SOP (min) | SPH (min) |
| --- | --- | --- | --- | --- | --- | --- | --- | --- | --- | --- |
| 2016 | Zhang & Parhi | MIT, 17 patients | power spectral density ratio | SVM | no | 80 | 98.68 | 0.05 | 50 | 0** |
| 2017 | Alotaiby et al | MIT, 23 patients | CSP | LDA | yes | 170 | 81 | 0.47 | 60 | 0 |
| 2018 | Khan et al | MIT, 15 patients | wavelet transform | CNN | yes | 18 | 83.33 | 0.15 | 10 | 0** |
| 2018 | Truong et al | MIT, 13 patients | short-time Fourier transform | CNN | yes | 64 | 81.2 | 0.16 | 30 | 5 |
| 2018 | Tsiouras et al | MIT, 23 patients | time domain, frequency domain, graph theory features, correlation features | LSTM | no | 185 | 100 | 0.06 | 30 | 0** |
| 2019 | Daoud & Bayoumi | MIT, 8 patients | DCAE + Bi-LSTM |  | yes | 43 | 99.6 | 0.004 | 60 | 0** |

\*\*SPH implicitly set to 0 in these works
Abbreviations: Features (Feats), Classifier (CLF), Seizures (Sz), Sensitivity (Sens), Same Feature engineering (FE), Same Calibration (Cal) denotes studies which were calibrated on the same dataset used for training.

## Notes

### Competing Interest Statement

The authors have declared no competing interest.

### Funding Statement

This study was funded by Regents of the University of California (Resource Allocation Program).

### Author Declarations

The source data used were publicly available before the initiation of the study and are available via the cited references.

